# Muddying the Waters: Syncytial Variant Nodular Sclerosis Classic Hodgkin Lymphomas Exhibit a Primary Mediastinal Large B-cell Lymphoma-Like Gene Expression Profile Using the Lymph3Cx Gene Expression Profiling Assay

**DOI:** 10.1101/2025.05.12.25327251

**Authors:** Jinjun Cheng, Sarah E. Gibson, Ryan Barry, Ryan S. Robetorye

## Abstract

Mediastinal B-cell lymphomas are relatively frequent in young patients and include nodular sclerosis classic Hodgkin lymphoma (NSCHL), primary mediastinal large B-cell lymphoma (PMBL), and rarely, mediastinal gray zone lymphoma (MGZL). Occasional NSCHLs contain abundant Hodgkin/Reed Sternberg (HRS) cells that exhibit a syncytial growth pattern, which may create diagnostic challenges. Previous studies have demonstrated that PMBL can be distinguished from subtypes of diffuse large B-cell lymphoma (DLBCL) based on gene expression signatures using the Lymph3Cx gene expression profiling assay, which has been validated as a clinical test in our molecular diagnostics laboratory. Here, we demonstrate that syncytial variant NSCHL exhibits a PMBL-like gene expression profile using the Lymph3Cx gene expression profiling assay. It is critical for pathologists and oncologists to be aware of this potential diagnostic pitfall to avoid possible misdiagnosis.

## Introduction

Primary mediastinal large B-cell lymphoma (PMBL), a thymic-B-cell origin lymphoma, shares pathologic, genetic, and clinical features with other B-cell lymphomas involving the thymus, such as classic Hodgkin lymphoma (CHL) and mediastinal gray zone lymphoma (MGZL).^1-3^ Currently, patients with these lymphomas may receive different treatments and have variable outcomes, despite the many similarities.^4,5^ In addition, current practice based on histopathology and clinical presentation may give rise to diagnostic inaccuracy. Therefore, establishing a consensus and evidence-based guideline for accurate classification is of critical importance for choosing optimal therapeutic strategies.

Multiple studies have shown that the gene expression profile (GEP), somatic mutations (e.g, *B2M* and *CD58* mutations), and chromosomal copy number alterations (e.g, rearrangements of *CIITA*, and abnormalities of 9p24.1) in PMBL resemble those of CHL and MGZL, but are distinct from conventional diffuse large B-cell lymphoma (DLBCL).^1-3,6-8^ Moreover, rare cases with similar GEP and/or genetic alterations reminiscent of PMBL have also been identified outside the mediastinum.^7-10^

Syncytial variant nodular sclerosis CHL (SVNSCHL), first described over 40 years ago, exhibits cohesive and confluent sheets of Hodgkin/Reed-Sternberg (HRS) cells, which may create diagnostic challenges.^11,12^ Although SVNSCHLs are uncommon, their clinical features overlap with those of PMBL, including a predominance in young adults and mediastinal mass presentation in over 90%.^13^ SVNSCHLs are also associated with poorer overall survival compared with non-SVNSCHL patients.^13^ To the best of our knowledge, there is an absence of available data comparing the clinical, pathological, GEP, and genomic changes between PMBL and SVNSCHL (both mediastinal and non-mediastinal). Although a prior study revealed variation of 43 genes between CHL and PMBL,^3^ it is unclear whether this variation might actually be due to the low number of HRS cells in the studied CHL cases.

## Methods and Results

A total of 28 cases of CHL were subjected to Lymph3Cx analysis in our clinical molecular diagnostic laboratory, including 10 cases of NSCHL, four cases of mixed cellularity CHL (MCCHL), two cases of lymphocyte rich CHL (LRCHL), one case of lymphocyte depleted CHL (LDCHL), and 11 cases of SVNSCHL (Table 1). All cases were reviewed by at least two hematopathologists (RSR, SEG, JC) who provided a consensus diagnosis. The diagnostic criteria that distinguish CHL from PMBL and MGZL were based on the current International Consensus Classification (ICC) and 5^th^ World Health Organization (WHO) classifications.^8,10^

**Table 1.** Summary of Classic Hodgkin Lymphoma Case Characteristics and Lymph3cx Analysis Results.

| Case # | Age range | Sex | Anatomic Location of Biopsy | Immunohistochemistry profile of HRS cells | Diagnosis | Lymph3Cx Assignment with Probability Score |
| --- | --- | --- | --- | --- | --- | --- |
| 1 | 41-50 | F | Cervical LN | CD30+, CD15+, CD45-, | SVNSCHL | PMBL (0.93) |
|  |  |  |  | CD20-, CD79a- |  |  |
| 2 | 31-40 | F | Mediastinum | CD30+, CD15+, CD45-,<br>cd20-, CD23+ (p,w),<br>CD200+, MAL1+ (p,w),<br>PDL1+, PDL2+ (p,w) | SVNSCHL | PMBL (0.99) |
| 3 | 21-30 | M | Axillary LN | CD30+, CD15+, CD45-,<br>PAX5+ (w), CD20-,<br>CD23+ (p,w), CD200+,<br>MAL1+ (w), PDL1+,<br>PDL2+ (p,w) | SVNSCHL | PMBL (0.99) |
| 4 | 21-30 | M | Cervical LN | CD30+, CD15+, CD45-,<br>CD79a-, PAX5+ (w),<br>CD20+ (p), CD23+,<br>CD200+(p), MAL1+ (w),<br>PDL1+, PDL2+ | SVNSCHL | PMBL (1.00) |
| 5 | 21-30 | F | Cervical LN | CD30+, CD15+, CD45-,<br>CD79a+(v), PAX5+ (w),<br>CD20+ (v), CD23+,<br>CD200+, MAL1+,<br>PDL1+, PDL2+(p,w) | SVNSCHL | PMBL (1.00) |
| 6 | 51-60 | M | Cervical LN | CD30+, CD15+, CD45-,<br>PAX5+ (w), CD20+ (r),<br>CD23+(p,w), CD200+,<br>MAL1+ (w), PDL1+,<br>PDL2+(p,w) | SVNSCHL | PMBL (0.99) |
| 7 | 51-60 | M | Axillary LN | CD30+, CD15-, CD45-,<br>PAX5+ (w), CD20- | SVNSCHL | PMBL (0.99) |
| 8 | 11-20 | F | Mediastinum | CD30+, CD15+, CD45-,<br>PAX5+ (w), CD20-,<br>CD23+(p,w), CD200<br>ND, MAL1+, PDL1+,<br>PDL2 ND | SVNSCHL | PMBL (1.00) |
| 9 | 41-50 | F | Axillary LN | CD30+, CD15+, CD45-,<br>PAX5+ (w), CD20+ (v),<br>CD23+(p), CD200 ND,<br>MAL1 ND, PDL1 ND,<br>PDL2 ND | SVNSCHL | PMBL (1.00) |
| 10 | 21-30 | M | Mediastinum | CD30+, CD15+, CD45-,<br>CD79a-, PAX5-, CD20-,<br>CD23+(r), CD200+,<br>MAL1+(w), PDL1+,<br>PDL2+(p, w) | SVNSCHL | PMBL (0.98) |
| 11 | 31-40 | F | Axillary LN | CD30+, CD15+, CD45-,<br>PAX5+ (w), CD20-,<br>CD23+(p), CD200+,<br>MAL1+(w), PDL1+,<br>PDL2- | SVNSCHL | PMBL (0.94) |
| 12 | 21-30 | M | Cervical LN | CD30+, CD15+, CD45- | NSCHL | UNCLEAR<br>(0.43) |
| 13 | 31-40 | M | Axillary LN | CD30+, CD15+, CD45- | NSCHL | UNCLEAR |
|  |  |  |  |  |  | (0.88) |
| 14 | 21-30 | M | Cervical LN | CD30+, CD15+, CD45-, CD20+ (p,w), PAX5+ (w) | NSCHL | UNCLEAR (0.47) |
| 15 | 61-70 | M | Supraclavicular LN | CD30+, CD15+, CD45-, PAX5+ (w), CD20+(r) | NSCHL | UNCLEAR (0.70) |
| 16 | 21-30 | F | Axillary LN | CD30+, CD15+, CD45-, PAX5+ (w), CD20- | NSCHL | UNCLEAR (0.25) |
| 17 | 21-30 | F | Cervical LN | CD30+, CD15-, CD45-, PAX5+ (w), CD20- | NSCHL | UNCLEAR (0.45) |
| 18 | 11-20 | F | Cervical LN | CD30+, CD15+, CD45-, PAX5+ (w), CD20- | NSCHL | DLBCL (0.07) |
| 19 | 61-70 | M | Peritoneal LN | CD30+, CD15+(w), CD45-, PAX5+ (w), CD20- | NSCHL | UNCLEAR (0.87) |
| 20 | 61-70 | M | Cervical LN | CD30+, CD15+, CD45-, PAX5+ (w), CD20- | NSCHL | UNCLEAR (0.60) |
| 21 | 31-40 | M | Mediastinum | CD30+, CD15+(w), CD45-, PAX5+ (w), CD20+(subset) | NSCHL | UNCLEAR (0.73) |
| 22 | 21-30 | M | Cervical LN | CD30+, CD15+, CD45-, PAX5+ (w), CD20+(v) | MCCHL | UNCLEAR (0.13) |
| 23 | 61-70 | F | Mediastinum | CD30+, CD15+, CD45-, PAX5+ (w), CD20- | MCCHL | DLBCL (0.07) |
| 24 | 11-20 | M | Cervical LN | CD30+, CD15+, CD45- | MCCHL | UNCLEAR (0.66) |
| 25 | 51-60 | M | Iliac LN | CD30+, CD15+, CD45- | MCCHL | UNCLEAR (0.33) |
| 26 | 61-70 | M | Mediastinum | CD30+, CD15+(w), CD45-, PAX5+, CD20+(subset) | LRCHL | DLBCL (0.01) |
| 27 | 71-80 | M | Supraclavicular LN | CD30+, CD15+, CD45-, PAX5+ (w), CD20- | LRCHL | DLBCL (0.02) |
| 28 | 51-60 | F | Cervical LN | CD30+, CD15+, CD45- | LDCHL | UNCLEAR (0.25) |

Abbreviations: HRS= Hodgkin Reed Sternberg cells; LN= lymph node; p= partial expression; v= variable expression; r= rare positive cells; ND= not done; NSCHL=nodular sclerosis classic Hodgkin lymphoma; MCCHL= mixed cellularity classic Hodgkin lymphoma; SVNSCHL = syncytial variant nodular sclerosis classic Hodgkin lymphoma; LDCHL = lymphocyte depleted classic Hodgkin lymphoma; PMBL= primary mediastinal large B-cell lymphoma; DLBCL= diffuse large B-cell lymphoma.

SVNSCHL was defined by many HRS cells with a cohesive apperance, and forming confluent sheets in many of the tumor nodules.^12,13^ The related radiology and clinical presentation were collected when available. This study was performed with Mayo Clinic Institutional Review Board approval under protocol #16-000507. The Lymph3Cx assay was performed as previously reported,^14^ and produces a calculated score on a scale of 0.00 to 1.00 to classify each sample based on the probability that the sample is PMBL.^14^ Samples with scores of 0.90-1.00 are classified as PMBL, those with scores of 0.00-0.10 are classified as DLBCL, and samples with scores >0.10 and <0.90 are categorized as UNCLEAR.

All 11 SVNSCHL cases (3 mediastinal and 8 non-mediastinal) analyzed by the Lymph3Cx assay showed a PMBL-like GEP with a PMBL probability score greater than 0.90, but none of the other CHL subtypes exhibited this GEP signature (Table 1). The most common gene expression result was UNCLEAR in the non-SVNSCHL cases. Interestingly, three CHLs, including one NSCHL, one MCCHL, and two LRCHL showed PMBL probability scores less than 0.10, most consistent with classification as DLBCL.

Immunohistochemical stains performed during the diagnostic work-up of all 28 CHL cases are listed in Table 1. Of note, all 11 cases of SVNSCHL were positive for CD15 and CD30, except for one case negative for CD15, and all 11 cases were negative for CD45. Evidence of B-cell lineage was also observed in most cases, with weak PAX5 staining in 10/11 cases, and rare CD20 positive cells or some evidence of partial/weak CD20 staining in four of 11 cases.

Importantly, there was no significant difference in the immunostaining pattern between the mediastinal SVNSCHL (n=3) and non-mediastinal SVNSCHL (n=8) lymphoma cases.

Additional immunostains were performed on nine of 11 SVNSCHL cases using various immunostains that are often found to be positive in cases of PMBL.^15^ All nine SVNSCHL cases showed variable expression of CD23, MAL, CD200, PDL1, or PDL2, ranging from partially and weakly positive to diffusely and strongly positive (Table 1).

Next generation sequencing (NGS) was performed on selected cases of SVNSCHL with sufficient remaining tissue (n=4), using a B-cell lymphoma panel containing 46 genes.

## Discussion

In summary, we analyzed 28 cases of CHL using the Lymph3Cx GEP assay and found that all 11 cases of SVNSCHL in our series exhibited a PMBL-like GEP signature, but none of the other CHL subtypes exhibited this signature. The SVNSCHL cases also expressed additional antigens commonly associated with PMBL, including CD23, MAL, CD200, PDL1 or PDL2.

These findings muddy the current differential diagnosis between CHL and PMBL cases. Interestingly, the SVNSCHL cases in our series exhibited a PMBL-like GEP signature regardless of mediastinal or non-mediastinal location. This phenomenon has also been described in other rare large B-cell lymphomas outside of the mediastinum.^7-10^

Importantly, it should be recognized that the Lymph3Cx assay in isolation cannot be used to discrimiate SVNSCHL from PMBL; instead, close evaluation of the histologic and immunophenotypic findings in combination with the clinical features remains essential for differentiating these two entities. The most important difference between PMBL and CHL remains the expession level of the B-cell program. In our study, we relied on CD45, CD20, PAX5, CD79a, CD30 and CD15 immunostaining to differentate SVNSCHL and PMBL.

However, “gray zone” lymphomas with ambiguous histological and immunophenotypic features of both PMBL and CHL are recognized, designated as MGZL when arising in the mediastinum.^16,17^ Such cases were not included in the current study, but will be further investigated by our group in the near future.

## Data Availability

All data produced in the present work are contained in the manuscript

## List of abbreviations

Abbreviation Full term

WHO: World Health Organization
ICC: International Consensus Classification
SVNSCHL: Syncytial variant nodular sclerosis classic Hodgkin lymphoma
PMBL: Primary mediastinal large B-cell lymphoma
DLBCL: Diffuse large B-cell lymphoma
MGZL: Mediastinal gray zone lymphoma
GEP: Gene expression profile

---

**Figure 1.**
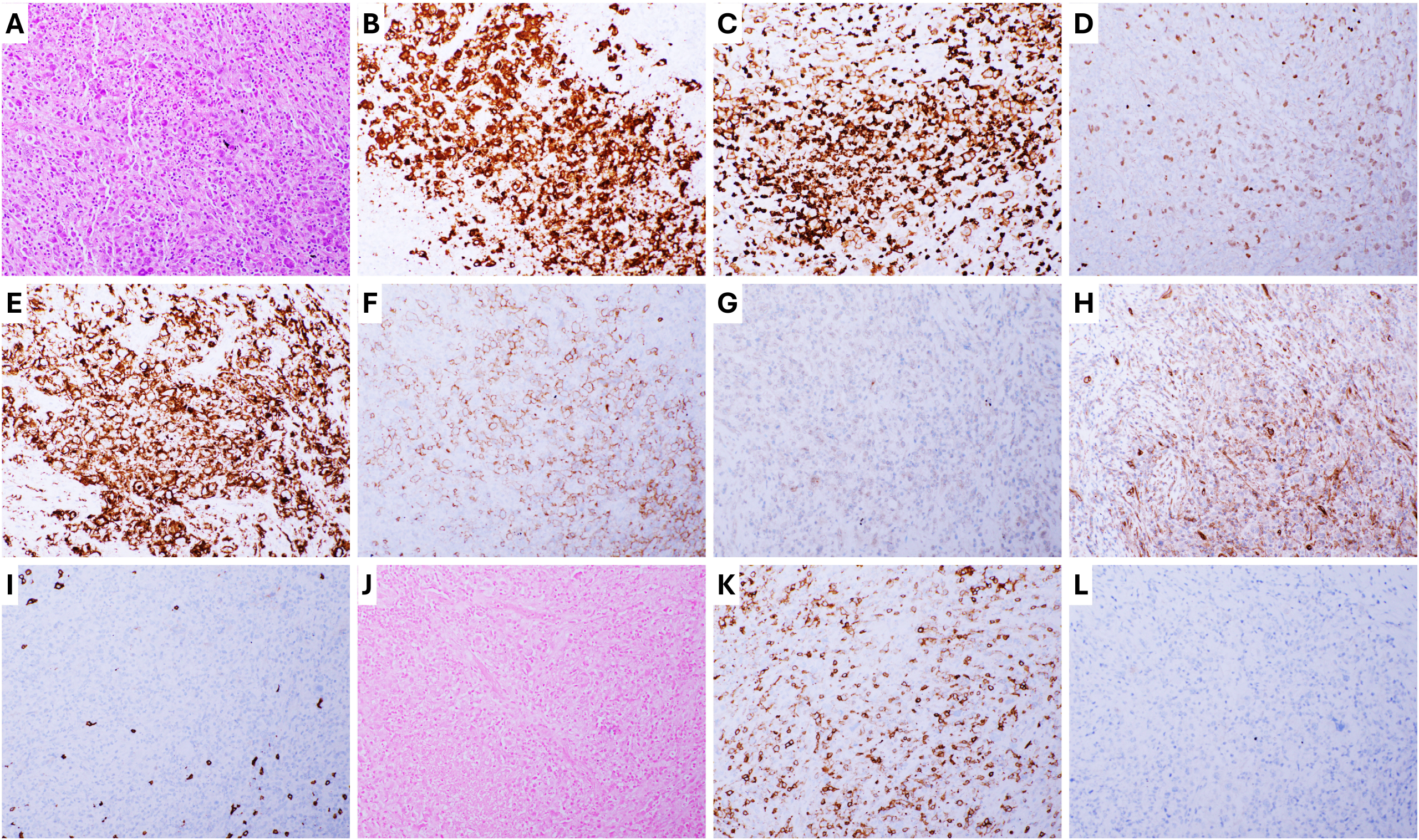
Representative case of syncytial variant nodular sclerosis classic Hodgkin lymphoma with Lymph3Cx PMBL-like gene expression signature. A, Hematoxylin and Eosin (H&E) staining, 200x, lymph node biopsy revealed sheets of Hodgkin Reed Sternberg (HRS) cells; The HRS cells are strongly positive for CD30 (B, 200x), CD15 (C, 200x), dim PAX5 (D, 200x), PDL1 (E, 200x), CD23 (F, 200x), partial and weak MAL1 (G, 200x), partial CD200 (H, 200x); negative with CD20 (I, 200x), EBER in situ hybridization (J, 200x), CD45 (K, 200x), and PDL2 (L, 200x).

